# Prospective study of the behavior of totally implantable long-term catheters in patients with mammary hypertrophy

**DOI:** 10.1101/2024.11.05.24315455

**Authors:** Maria Fernanda C. Portugal, Bruno Jeronimo Ponte, Carolina Carvalho Jansen Sorbello, Thulio Fernandes de Souza, Amanda Indig Pinheiro, Andressa Cristina Sposato Louzada, Dafne Braga Diamante Leiderman, Nelson Wolosker, Cynthia de Almeida Mendes

## Abstract

**Background:** The aim of this study was to evaluate the change in the position of totally implantable venous catheters (TIVC) tips in patients with mammary hypertrophy, comparing them to patients without this condition in both the supine and upright position.

**Methods:** This was a prospective study involving consecutive patients who underwent the implantation of totally implantable central venous catheters (TIVC) in internal jugular or subclavian veins with reservoir placement on the thoracic wall. Data were collected with regard to demographic and clinical features of the included patients, including: age (years), height (cm), weight (Kg), BMI, malignancy type, occurrence of previous non-deforming procedures to the breasts, presence of a Peripherally Inserted Central Catheter (PICC) during TIVC implantation, and history of DVT (punctured vein and laterality). The catheter tip’s position in an upright stance was documented through a routine postoperative chest X-ray, using a posterior-to-anterior incidence technique, also including a radiopaque ruler permitting the measuring of its distance from the carina in centimetres. The intraclass correlation coefficient between the intraoperative and postoperative distance variation recorded by Observers 1 and 2 was excellent (r =0.773).

**Results:** Significant caudal TIVC tip displacement was determined by previous nondeforming surgery to the chest wall (p=0.002); and significant proximal displacement was calculated by the presence of PICC during TIVC implantation (p =0.042). The Sacchini index did not exhibit a significant association with changes in catheter tip locations after surgery.

**Conclusion:** Significant caudal TIVC tip displacement was determined by previous non-deforming surgery to the thoracic wall (p=0.002); and significant proximal displacement was determined by the presence of a PICC during TIVC implantation (p=0.042). Mammary hypertrophy, as assessed by the Sacchini index, was not statistically correlated to catheter tip displacement (p=0.612).

## Introduction

Long-term totally implantable central venous catheters (TIVC) are extensively used in medical practice, primarily for patients who require frequent venous access, particularly for administering potent or unsuitable drugs via peripheral routes. These catheters are commonly recommended for oncology patients undergoing chemotherapy. (1–3)

For these devices to function properly, it is essential that they are positioned correctly and that there is no deformation during their use. (4)In order to study the dynamics of catheter positioning, Vesely et al., in 2003, studied central catheters of peripheral insertion, identifying two anatomical factors in the movement of the catheter tips for large-diameter catheters in subclavian or internal jugular veins. (5) Firstly, the shift in abdominal contents from the supine to the upright position elongates central veins, impacting catheter tip placement near the right atrium. Additionally, the caudal movement of the thoracic wall in an upright posture causes a cranial shift in the catheter tip, pulling it away from the cavo-atrial junction, the ideal position. (5)

Studies assessing catheter movement in these conditions have reported changes of up to 3.2 cm(6), with greater effects observed in women and obese patients, particularly for larger catheters. (7)

Mammary hypertrophy is a common condition in women and occurs when there is an increase in glandular tissue during puberty or an increase in the deposition of adipose tissue. This leads to an increase in the weight of the breast, causing greater caudal retraction.(8) Thus, many believe, without scientific basis, that patients with breast hypertrophy who are undergoing TIVC implantation are considered to be at greater risk of catheter tip migration due to the effects of thoracic wall forces in the upright position.

This study aims to evaluate the change in the position of TIVC tips in patients with mammary hypertrophy, comparing them to patients without this condition in both the supine and upright position.

## Methods

### Study design and patient enrolment

This prospective study involved consecutive patients who underwent the implantation of fully implantable central venous catheters (TIVC) at a Cancer hospital of tertiary complexity from August 2020 to August 2022.

This study adhered to the principles outlined in the Helsinki Declaration and received approval from the Ethics Committee of the Hospital Israelita Albert Einstein under protocol number 45305121.5.0000.0071. Before participation, all patients received comprehensive information about the study’s objectives and procedures, and informed consent was obtained from each participant.

### All data was handled confidentially and de-identified

Patients were included if they were over 18 years of age, female, with a formal indication for the implantation of these devices by their attending medical teams and if they consented to participate by signing an Informed Consent Form. The sample included patients who underwent the implantation of TIVCs in internal jugular or subclavian veins with reservoir placement on the thoracic wall.

Patients who could not tolerate chest X-rays in an upright position, those experiencing intraoperative complications, significant hematomas, genetic or mass-induced anatomical alterations, prior surgeries incurring anatomical distortion, or patients with reservoirs placed in locations other than the anterior thoracic wall were excluded.

### Data collection and subgroup analysis

Data was collected about demographic and clinical features of the included patients, including age (years), height (cm), weight (Kg), BMI, malignancy type, occurrence of previous non-deforming procedures to the breasts or thoracic wall, presence of a Peripherally Inserted Central Catheter (PICC) at the time of TIVC implantation, history of a previous catheter in the implantation site; history of DVT in the implantation site and implantation site (punctured vein and laterality).

Patients were categorized into two groups based on breast volume, distinguishing between those with breast hypertrophy and the control group. The assessment relied on the Sacchini index, which was obtained preoperatively in the surgical suite with the patient in an upright position.

The Sacchini index is a measurement based on the proportion of breast dimensions taken with a measuring tape, eliminating the need for invasive procedures (9). Two measurements are taken: from the lateral edge of the sternum to the nipple and from the inframammary fold to the nipple. The index is calculated as the average of these measurements. Breasts with measurements between 9 and 11 cm are classified as average in size, while those with measurements exceeding 11 cm are considered hypertrophic, with each breast measured individually. (10) The patients were assigned to one of two groups based on the Sacchini index result on the side corresponding to the planned device insertion.

### Procedure technique

The institutional protocol determines implantation via a surgical procedure in an operating theatre, with the patient in a supine position under sedation and local anesthesia. The venous puncture was always ultrasound-guided, and catheter implantation was always performed under fluoroscopy.

The Bard Port (Bard Access Systems, INC, Salt Lake City, Utah, USA) device was used for all procedures.

The venous implantation site was decided upon bedside ultrasound evaluation, considering vein diameter, compressibility, and phasicity, in this preference order: right internal jugular vein, left internal jugular vein, right subclavian vein, and left subclavian vein.

### Intraoperative Catheter Tip Position Documentation

The immediate postoperative position of the catheter tip was recorded using a fluoroscopic image at the end of the procedure and noted using a radiopaque ruler placed on the surgical table to measure its distance from the carina in centimeters (Figure 1).

**Figure 1:**
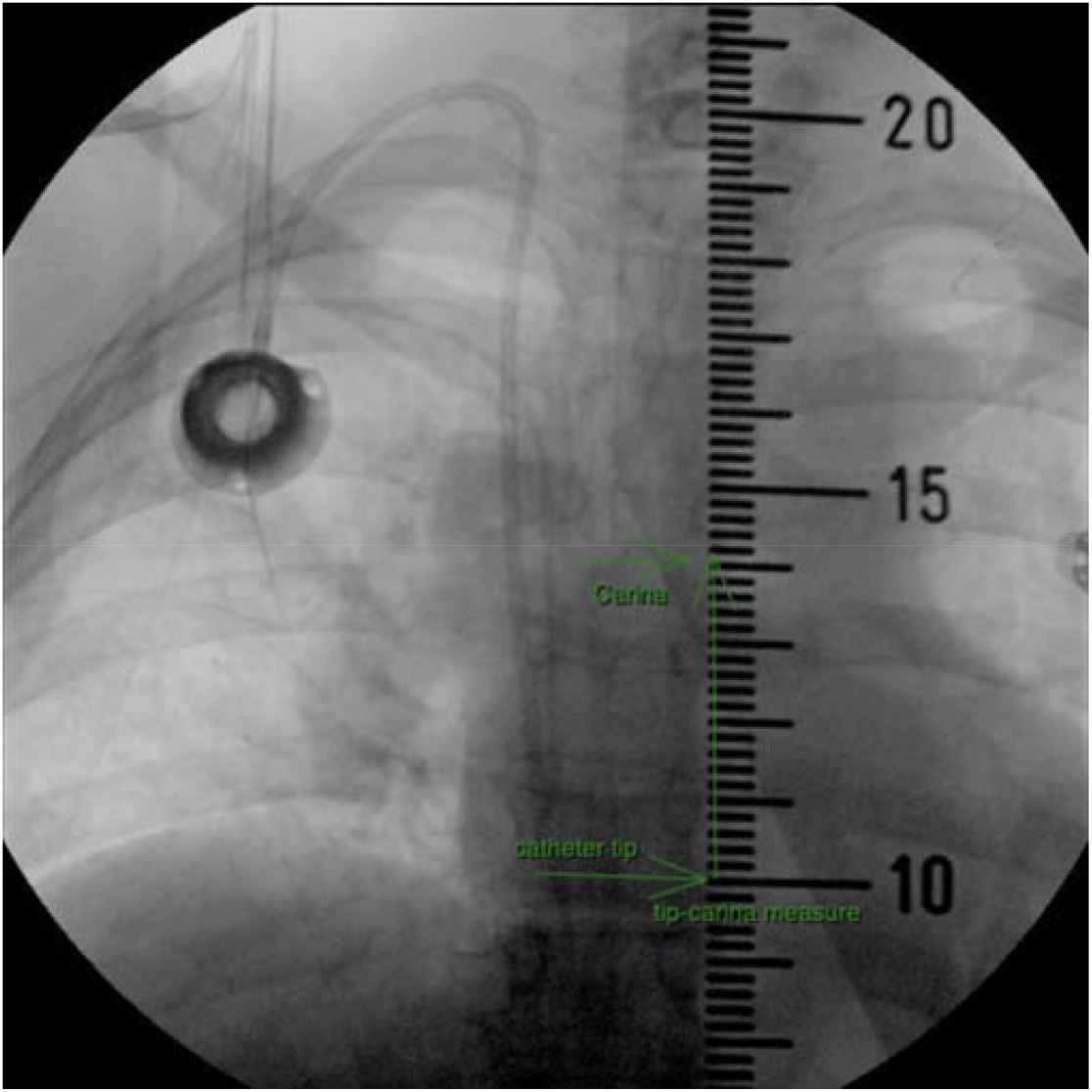
Intraoperative image of the measure of the catheter tip distance to the carina.

### Postoperative Upright Catheter Tip Evaluation

The catheter tip’s position in an upright stance was documented through a routine postoperative chest X-ray, using a posterior-to-anterior incidence technique, also including a radiopaque ruler permitting the measuring of its distance from the carina in centimeters (Figure 2).

**Figure 2.**
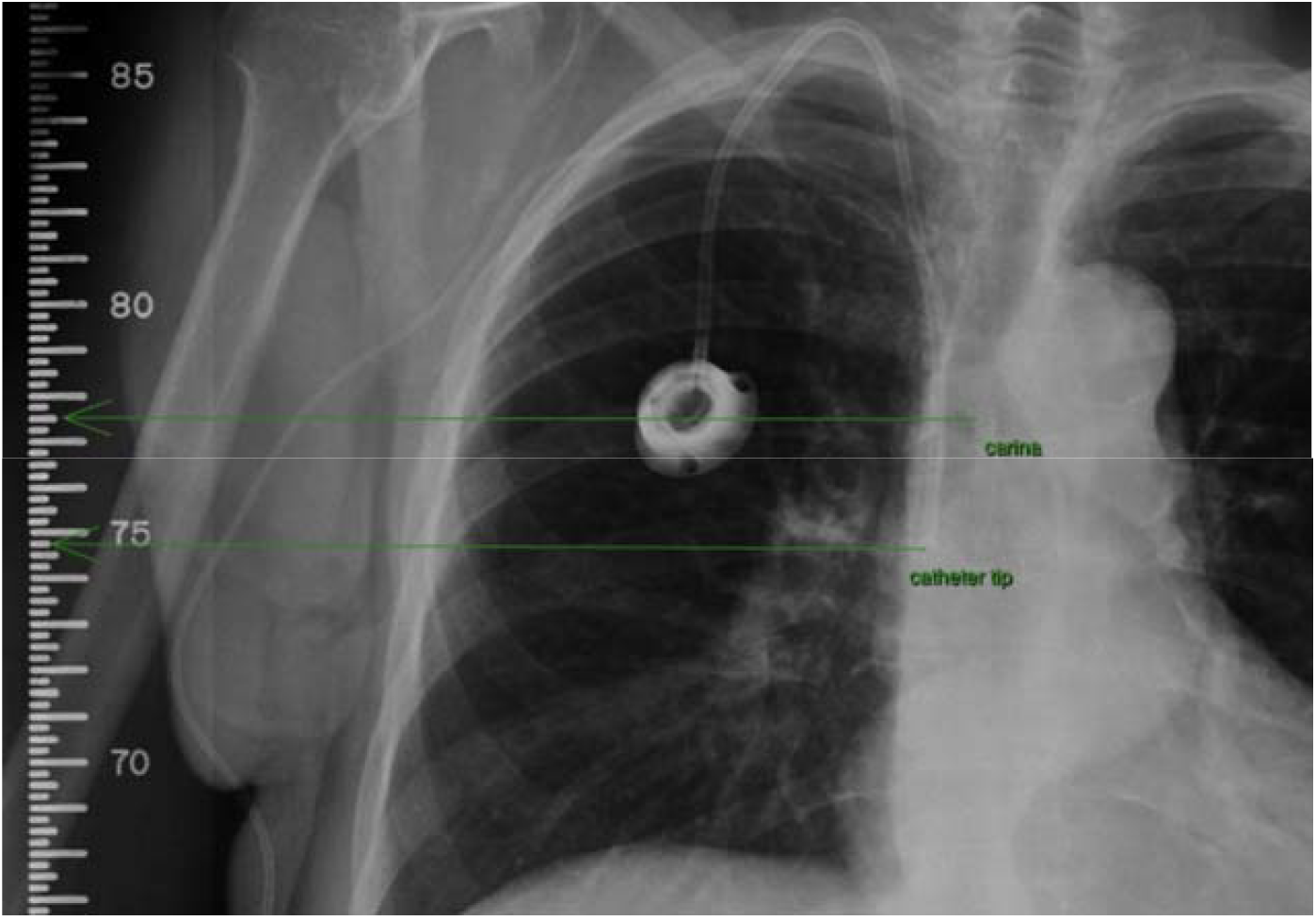
Postoperative standing X-ray image of the measure of the catheter tip distance to the carina

### Data analysis and interobserver validation

The demographic data of the two groups of patients were collected and analyzed.

Two observers and team members responsible for the procedures were selected to measure the distances between the catheter tip and the carina during the intraoperative (horizontal decubitus) and postoperative periods (orthostasis).

Intraoperatively, a radiopaque ruler was positioned below the patient to take the measurement using radioscopy. Postoperatively, the patient was referred for a chest X-ray in orthostasis, a standard procedure for patients undergoing catheter implantation at the hospital. The variation in the positioning of the catheter tip was expressed in millimeters.

Negative values represented an approximation to the carina (shortening), while positive values represented a distancing from the carina (lengthening). For analysis purposes, the mean value of the tip variation between the breast hyperplasia and control groups was calculated and compared.

Observer 2’s annotations aimed to validate those made by Observer 1 using the Intraclass Correlation Coefficient. Observer 1’s recorded distances served as the benchmark for all statistical analyses

### Statistical Analysis

The data’s normal distribution was confirmed through the Shapiro-Wilk test. Demographic and clinical characteristics were presented as relative frequencies and mean values with their respective standard deviations.

Qualitative characteristics related to TIVC tip displacement were evaluated using the Student’s T-test, while quantitative characteristics were assessed in terms of TIVC tip displacement using Pearson’s correlation.

All statistical analyses were conducted using the SPSS software, version 25.0. Significance level was set at p<0.05.

## Results

A total of 116 patients were included. Patient enrolment is detailed in Diagram 1.

**Diagram 1:**
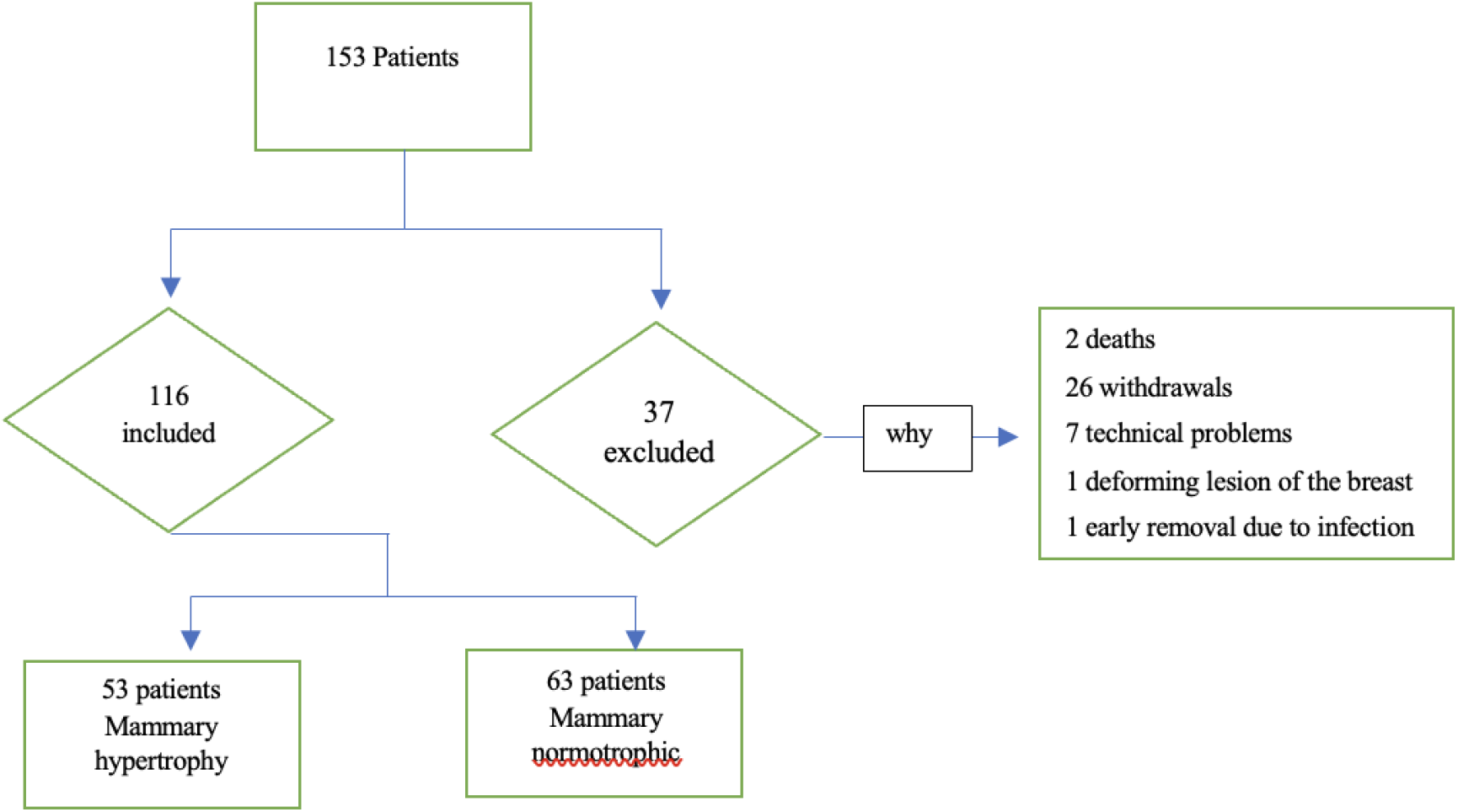
Patient enrolment flowchart

Demographic and clinical characteristics of the sample are presented in **Table 1**.

**Table 1:** Demographic and characteristics of patients.

| Variable | *n* / average | (%) / SD |
| --- | --- | --- |
| Age (Years) | 57,52 | ±15,48 |
| Weight | 63,80kg | ±17,87kg |
| Lenght | 157,56cm | ±7,16cm |
| BMI | 25,59kg/m<sup>2</sup> | ±6,5kg/m<sup>2</sup> |
| Sacchini Index | 107,17mm | ±30,13mm |
| Neoplasia |  |  |
| Gastrointestinal | 67 | 57,76% |
| Hemotologic | 18 | 15,52% |
| Breast | 16 | 13,79% |
| Genitourinary | 8 | 6,90% |
| Lung | 4 | 3,45% |
| Musculoskeletal | 3 | 2,59% |

*SD*: Standard deviation; *BMI*: body mass index

The Intraclass Correlation Coefficient between the intraoperative and postoperative distance variation recorded by Observers 1 and 2 was excellent (*r* =0.773) (11), and is described in Table 2.

**Table 2:** Interobserver concordance for the catheter tip displacement.

| Variable | N | Average (mm) | SD | ICC | IC (95%) |  | Repeatability |
| --- | --- | --- | --- | --- | --- | --- | --- |
|  |  |  |  |  | Lower | Upper |  |
| Carina distance – catheter tip intraoperative (mm) OBS 1 | 116 | 47,1 | 14,5 | 0,853 | 0,795 | 0,896 | 5,71 |
| Carina distance – catheter tip intraoperative (mm) OBS 2 | 116 | 46,8 | 15,2 |  |  |  |  |
| Carina distance – catheter tip orthostasis (mm) OBS 1 | 116 | 48,4 | 20,6 | 0,926 | 0,885 | 0,952 | 5,13 |
| Carina distance – catheter tip orthostasis (mm) OBS 2 | 116 | 45,9 | 19,1 |  |  |  |  |
| Variation OBS 1 | 116 | -1,32 | 15,36 | 0,712 | 0,607 | 0,792 | 7,73 |
| Variation OBS 2 | 116 | 0,93 | 13,73 |  |  |  |  |

*ICC: Intraclass correlation coefficient; SD: Standard deviation; IC: interval of confidence; OBS: observer*

To catheter tip movement, as detailed in Table 3, significant displacement was only determined by previous non-deforming surgery to the thoracic wall (*p*=0.002) and the presence of a PICC during TIVC implantation (*p*=0.042). Mammary hypertrophy did not correlate with tip displacement during follow–up (p 0,182).

**Table 3:** Patients characteristics and correlation with tip displacement.

| Qualitative Characteristics | N | Average (mm) | Standard Deviation | Median | Minimum | Maximum | p-value |
| --- | --- | --- | --- | --- | --- | --- | --- |
| **Previous surgery to the thoracic wall** |  |  |  |  |  |  | **0,002\*** |
| No | 112 | -0,52 | 14,79 | 1,5 | -38 | 31 |  |
| Yes | 4 | -23,88 | 15,83 | -24,75 | -40 | -6 |  |
| **Presence of a PICC during procedure** |  |  |  |  |  |  | **0,042\*** |
| No | 85 | -2,89 | 16,11 | -5 | -40 | 29 |  |
| Yes | 31 | 2,97 | 12,33 | 3 | -18,5 | 31 |  |
| **Previous catheter in the implanted vessel** |  |  |  |  |  |  | 0,357* |
| No | 103 | -0,85 | 15,29 | 0 | -40 | 31 |  |
| Yes | 13 | -5,04 | 16,05 | -1 | -36 | 13,5 |  |
| **Previous thrombosis on the implanted side** |  |  |  |  |  |  | 0,167* |
| No | 115 | -1,14 | 15,30 | 0 | -40 | 31 |  |
| Yes | 1 | -22,50 | 0,00 | -22,5 | -22,5 | -22,5 |  |
| **Chest mass that causes deformity** |  |  |  |  |  |  | 0,559* |
| No | 110 | -1,13 | 14,86 | 0 | -38 | 31 |  |
| Yes | 6 | -4,92 | 24,37 | -1 | -40 | 20 |  |
| **Port implante side** |  |  |  |  |  |  | 0,923* |
| Left | 8 | -0,81 | 16,27 | 2 | -30 | 20 |  |
| Right | 108 | -1,36 | 15,37 | 0 | -40 | 31 |  |
| **Group** |  |  |  |  |  |  | 0,182* |
| Mammary hypertrophy | 53 | -3,41 | 16,16 | -5 | -40 | 27 |  |
| Mammary normotrophic | 63 | 0,43 | 14,55 | 2 | -36 | 31 |  |
| **Quantitative Characteristics** |  |  | **Correlation** |  |  | **p-value** |  |
| BMI |  |  | - 0,026 |  |  | 0,779<sup>#</sup> |  |
| Age (Years) |  |  | 0,121 |  |  | 0,195<sup>#</sup> |  |
| Sacchini Index |  |  | -0,048 |  |  | 0,612<sup>#</sup> |  |

\*:Student T-test; #: Pearson correlation; BMI: body mass index

Mammary hypertrophy, as assessed by the Sacchini index, was not statistically correlated to catheter tip displacement (*p*=0.612).

## Discussion

The use of TIVC is widespread in patients who require long-term intravenous treatment. Properly installed and managed, TIVCs prove reliable and convenient for the extended treatment of malignancies. (2) The technique for their installation varies by center and patient condition, but generally, the catheter’s distal portion should be positioned in the superior vena cava at the atrio-caval junction. The reservoir should be located on a stable surface that permits easy access and management(4), with the anterior thoracic wall, particularly in the deltopectoral groove, being the preferred site. (1,12)

The functionality of fully implantable catheters, specifically their ability to infuse drugs and aspirate blood effortlessly, depends on the proper placement of the reservoir, the catheter tip, and the subcutaneous path. (1,13) Despite their undeniable usefulness, TIVCs can develop complications that can seriously affect the prognosis and quality of life in cancer patients. (1,2,14) Catheter thrombosis and infection are the two main complications related to these devices, with migration of the catheter tip acting as a significant risk factor for thrombosis and device malfunction. (14–16)

According to Vesely et al., in the supine position, mediastinal structures, including central veins, face compression due to abdominal contents. (5) In the upright position, abdominal contents shift caudally, leading to the elongation of central veins and an expansion of the right atrium. Given that the proximal catheter segment remains fixed (subcutaneous reservoir), this stretching of mediastinal structures may alter the position of the distal catheter tip concerning the superior vena cava and the right atrium. (5)

Another anatomical factor influencing the final position of catheter tips pertains to their tunnelling and implantation in a subcutaneous reservoir on the anterior thoracic wall. In the upright position, the thoracic wall moves caudally under the influence of gravity. The sutured proximal catheter segment follows this movement, pulling the intravascular segment of the distal tip cranially within the mediastinum and affecting the catheter’s tip position. Notably, in the upright position, the catheter tip tends to shift cranially away from the cavo-atrial junction, its ideal position. (5–7)

Contrary to expectations regarding breast volume, our analysis revealed that prior breast surgery and the presence of PICC catheters at the time of TIVC implantation had a statistically significant impact on postoperative catheter tip displacement. Previous breast surgery resulted in an average caudal displacement of 23.88 mm, which may be associated with anatomical breast changes (altering the volume-weight ratio) or scar tissue retractions in the thoracic wall. (5–7)

Conversely, the presence of PICC catheters during implantation led to an average cranial progression of 2.97 mm, potentially due to mechanical conflicts in the vascular lumen or device manipulation, resulting in both displacements. (17)

In contrast, breast volume, as assessed by the Sacchini index, did not exhibit a significant association with changes in catheter tip locations after surgery. This finding might be limited by factors such as breast density, which could lead to gravitational impacts of the breast on the thoracic wall, not necessarily linked to its volume.

While our study provides valuable insights into the factors influencing catheter tip displacement following TIVC implantation, it is essential to acknowledge several potential limitations. Firstly, our research was conducted within a specific patient population and clinical setting, and these findings may not be directly generalizable to other healthcare environments or patient demographics. Additionally, we should recognize that the sample size, while sufficient for our primary analyses, could influence the ability to detect more nuanced associations. Furthermore, the impact of unmeasured confounding variables and the potential for interobserver variability in distance measurements should be considered. Finally, while our results reveal statistically significant associations, the clinical relevance and broader implications of catheter tip displacement require further investigation and validation through prospective studies.

Despite its limitations, this study contributes valuable insights into a previously unexplored issue in clinical practice. The findings offer a foundation for further research and may serve as a practical tool in guiding future TIVC implantation procedures. By identifying significant factors associated with catheter tip displacement – prior non-deforming thoracic wall surgery and the presence of a PICC at the time of TIVC implantation –this study enhances our understanding of potential risk factors and allows for more informed decision-making in clinical settings.

## Conclusion

Significant caudal TIVC tip displacement was determined by previous non-deforming surgery to the thoracic wall (*p*=0.002), and significant proximal displacement was determined by the presence of a PICC during TIVC implantation (*p*=0.042).

Mammary hypertrophy, as assessed by the Sacchini index, was not statistically correlated to catheter tip displacement (*p*=0.612).

## Data Availability

All data produced in the present study are available upon reasonable request to the authors.

## Notes

### Competing Interest Statement

The authors have declared no competing interest.

### Funding Statement

This study did not receive any funding

### Author Declarations

Ethics committee / IRB of Hospital Israelita Albert Einstein gave ethical approval for this work.

